# Performance Decay of Molecular Assays Near the Limit of Detection: Probabilistic Modeling using Real-World COVID-19 Data

**DOI:** 10.1101/2021.04.26.21254638

**Authors:** Thomas J.S. Durant, Christopher D. Koch, Christopher A. Kerantzas, David R. Peaper

**Affiliations:** Yale University School of Medicine, Department of Laboratory Medicine: 55 Park Street PS345D, New Haven, CT 06511

**Keywords:** Viral burden, cycle threshold, sensitivity, detection rate, sars-cov-2, covid-19, bootstrap resampling

## Abstract

The gold standard for diagnosis of COVID-19 is detection of SARS-CoV-2 RNA by RT-PCR. However, the effect of systematic changes in specimen viral burden on the overall assay performance is not quantitatively described. We observed decreased viral burdens in our testing population as the pandemic progressed, with median sample Ct values increasing from 22.7 to 32.8 from weeks 14 and 20, respectively. We developed a method using computer simulations to quantify the implications of variable SARS-CoV-2 viral burden on observed assay performance. We found that overall decreasing viral burden can have profound effects on assay detection rates. When real-world Ct values were used as source data in a bootstrap resampling simulation, the sensitivity of the same hypothetical assay decreased from 97.59 (95% CI 97.3-97.9) in week 12, to 74.42 (95% CI 73.9-75) in week 20. Furthermore, simulated assays with a 3-fold or 10-fold reduced sensitivity would both appear to be >95% sensitive early in the pandemic, but sensitivity would fall to 85.55 (95% CI 84.9-86.2) and 74.38 (95% CI 73.6-75.1) later in the pandemic, respectively. Our modeling approach can be used to better quantitate the impact that specimen viral burden may have on the clinical application of tests and specimens.

## INTRODUCTION

On February 4, 2020, the U.S. Department of Health and Human Services (HHS) declared a public health emergency and authorized the emergency use of in vitro diagnostics (IVD) for the detection of the severe acute respiratory coronavirus 2 (SARS-CoV-2). Since this time, the number of emergency use authorizations (EUA) issued by the FDA, and the number of NAATs available for clinical use, have increased linearly. Currently, the required data for EUA are primarily those which describe the analytic sensitivity and specificity, and among available assays, these performance metrics are generally well described. The FDA only recently released (https://bit.ly/2RUL3R2) results of testing using a standardized panel of contrived viral samples, and a group recently compared assay limits of detection (LOD) using a quantitated set of viral standards.(1) Finally, there have been several method to method comparisons of SARS-CoV-2 testing.(1–14) However, at the time of this writing, the clinical performance of real-time reverse transcription-PCR (RT-PCR) based assays remains a difficult metric to evaluate.

The preferred method for a laboratory diagnosis of acute COVID-19 is an amplified RNA test, and RT-PCR is the most commonly used method. Among published reports of suspected clinical inaccuracies observed with SARS-CoV-2 RT-PCR, both false positive and false negative results have been reported. False positive results can occur due to specimen contamination, but analytical false positives have been reported to the FDA. Much more concern has been focused on falsely negative results driven by variation in test sensitivity. There are many variables in the testing process which arguably influence the diagnostic sensitivity of RT-PCR-based assays. Preanalytical conditions which are often cited include, timing of sample collection relative to the duration of illness, severity of illness, sample collection site (e.g. nasopharyngeal swab (NPS), nasal swab), and sample collection technique.(15, 16) Collectively, these variables directly influence viral burden in clinical samples, and can therefore influence diagnostic test performance. From an analytical standpoint, as the viral burden in clinical samples approaches or passes the LOD, the probability of qualitative detection decreases. As a result, specimen viral burden is a key component of overall test performance, both in the analytic and clinical aspects of assay consideration.

The diagnostic sensitivity of a test method is thought to be fairly constant across time, but publications evaluating the performance of different COVID-19 diagnostic assays have demonstrated highly variable sensitivity even for commercial assays with constant procedures. Additionally, studies of different specimen types with potentially different viral burdens have not been consistent in their reported sensitivity.(15–18) This has profound implications for clinical decision making since different levels of assay sensitivity encompassing specimen, collector, and assay may be required for different clinical scenarios.

As of December 2020, there is no commonly available and recognized reference material for quantitative viral load testing for SARS-CoV-2. Accordingly, the cycle threshold (Ct) value of real time RT-PCR assays, when available, is the only widely available semi-quantitative indicator of viral burden, and Ct value is inversely proportional to viral burden in a given sample. The Ct value represents the number of completed PCR amplification cycles it takes for the fluorescent signal, denoting the target gene, to increase beyond a threshold for positivity. Limitations of the Ct value include an inability to generalize between platforms, within-platform variability between laboratories, and the absence of available Ct values from some IVD instruments and analytical methods. Despite these limitations, using the Ct value to evaluate viral burden in clinical samples is a common clinical practice, and is often discussed in COVID-related literature as having potential as a prognostic indicator. In addition, it is often requested by clinicians to help explain discrepancies between provider clinical impression and a positive test result. However, there is limited evidence to support the widespread use of Ct values in these circumstances.

From an analytic standpoint, the Ct value remains a primary determinant in producing a qualitative result. In addition, there are also emerging reports on how Ct values of the underlying population may influence the diagnostic performance of an assay. In a recent study by Green et al., Ct values reported from samples where SARS-CoV-2 RNA was detected and stratified by patients who received repeat testing versus a single test. Testing was performed on the Cobas 6800 assay, and between single and repeat testing cohorts, they observed 3.38 and 6.59% increases in Ct values for gene targets 1 and 2, respectively.(10) Accordingly, the authors note that this difference in viral burden can influence the clinical sensitivity of the assay. While there are emerging reports which discuss the effect of variable viral burden in samples with respect to patient-level diagnosis, the degree to which shifting Ct values may influence the diagnostic sensitivity has not been quantitatively described.

During the initial weeks of the pandemic, we observed increasing rates of false negative results during validation experiments at our institution. Upon investigation we noted higher Ct values among the false negative samples. While it is known that higher Ct values are less likely to be detected, there was a higher proportion of low viral burden specimens in our sample cohort relative to the beginning of the pandemic. Given the potential impact of viral burden on test positivity, we sought to quantitate the systematic distribution of Ct values in the tested population from our institution across epidemiologic weeks. Further, because it was not practical to do multiple comparisons across multiple platforms to reliably describe differences in assay performance due to changes in sample viral burden, we employed mathematical modeling to quantitatively describe this. To this end, we used publicly available data to develop a model of test performance characteristics corresponding to different viral burdens allowing probabilistic-based simulation of RNA detection for a hypothetical COVID-19 assay evaluated at different timepoints during the pandemic. We then developed a computational model to simulate the assessment of RT-PCR clinical performance at our institution, using a bootstrap resampling method to describe variations in test sensitivity based upon different underlying assay performance assumptions. We hypothesize that mathematical modeling will demonstrate a temporal decay in diagnostic sensitivity as viral burden in clinical samples decreased.

## METHODS

### SARS-CoV-2 RNA Molecular Testing

Multiple molecular-based methods were used for detecting SARS-CoV-2 RNA during the observation period of this project. The platforms which performed more than 95% of testing within our healthcare delivery organization (HDO) included (1) the Panther Aptima SARS-CoV-2 Assay (Hologic; San Diego, CA) (2) the TaqPath COVID-19 Combo Kit (Thermo Fisher Scientific, Inc; Carlsbad, CA), (3) the Xpert Xpress SARS-CoV-2 test (Cepheid; Sunnyvale, CA), and (4) a modified version of the CDC assay. The Xpert Xpress and Aptima SARS-CoV-2 testing were performed at different sites within a 5-hospital HDO in Connecticut and Rhode Island starting March 28, 2020 and May 15, 2020, respectively. All testing was performed within clinical laboratories consistent with the manufacturer’s instructions. The modified CDC assay was performed at a single laboratory within our HDO beginning on March 13, 2020. It was granted Emergency Use Authorization by the US FDA, and has been previously described.(2) The preferred specimen type across all platforms was a NPS collected in 3 mL of universal or viral transport medium (VTM), but other specimen types and collection methods were used as needed based upon supply and reagent availability.

### Data Collection and Analysis

Retrospective laboratory test order and result data were extracted from our electronic health record (EHR) and laboratory information system (LIS). Row-level data, indexed by container ID, included test result interpretation (e.g., ‘detected’, ‘not detected’, etc.), Ct value, test method, and a collection datetime stamp. Additional metadata included patient class (e.g., inpatient, outpatient, or emergency department) at the time of their encounter, specimen collection type, and the resulting laboratory.

Custom scripts for data processing and analysis were implemented in Python (version 3.7.4), and R (version 4.0.2). Linear regression of Ct values associated with each gene target was performed in GraphPad Prism (version 8.4.2) (GraphPad Software; San Diego, CA, USA). Ct value analysis was limited to the platforms which provide a Ct value for each gene target which included the modified CDC assay and the Xpert assay. Tests which were performed at reference laboratories – i.e., ‘send-out’ tests – were not included in the Ct value analysis. CDC epidemiologic week numbers are used throughout the entirety of this report.

### Standardized assay performance analysis

Instructions for use (IFU) available on the US FDA website were reviewed on August 13, 2020. Limit of detection performance data were extracted from IFU if they met the following criteria: 1) The test was commercially available, 2) the test was an extracted RT-PCR assay, 3) at least 20 samples were tested at each concentration. Four assays met these criteria: Roche Cobas SARS-CoV-2, Abbott Alinity m SARS-CoV-2 (Abbott Molecular Inc; Des Plaines, IL), Cepheid Xpert Xpress SARS-CoV-2, and Perkin Elmer New Coronavirus Nucleic Acid Detection Kit (Perkin Elmer; Waltham, MA). The LOD, concentration tested, number of samples tested, and number of samples detected were recorded for each assay. The data from the Roche and Perkin Elmer assays were presented by RT-PCR target, while the data for the Abbott and Cepheid assays were presented for the total assay. For the purposes of calculations and curve fitting, the LOD of the Cepheid assay was changed to 0.005 PFU/mL from the claimed LOD of 0.02 PFU/mL since 95% of samples tested were positive at 0.005 PFU/mL. This revised LOD was consistent with probit analysis determined LOD (Data Not Shown). To control for different LODs and units of measure, each concentration tested was converted to a %-LOD. Individual samples with %-LOD greater than 250% were excluded from further analysis. The %-Detected was calculated for each assay or gene target. The %-Detected was plotted versus the %-LOD, and non-linear regression analysis with an exponential plateau function (where; Y_min_ = 0 and Y_max_ = 100) was fit in GraphPad Prism. Additionally, the fraction detected was used to determine a probit value, probit versus Log(10) %-LOD was plotted, and linear regression was performed (GraphPad Prism v8).

### Test Performance Simulation Model

The purpose of the model simulation was to determine – for a given cohort of positive samples – what fraction would be detected given the viral burden in the sample, and across the tested cohort, what is the calculated sensitivity. To this end, a function to calculate the probability of detection, on a per sample basis, was extrapolated from the non-linear regression model of the IFU data described above. Clinical performance of a hypothetical test was first simulated using Ct values from the modified CDC assay, pertaining to the N2 gene target. All N2 Ct values across the observational period, which were derived from a sample that was clinically reported as ‘detected’, were collected from the LIS (Figure 1A). These were subsequently collated into arrays and separated into groups by epidemiologic week (Figure 1B). From each dataset, corresponding to an epidemiology week, individual samples and their corresponding Ct value were randomly selected using a bootstrap resampling method with replacement (Figure 1C).(19) The Ct value was then used as input for the probability function (*P*) (Figure 1D). The Ct value at the LOD (Ct_LOD_) for the hypothetical assay for this simulation was defined as the viral concentration corresponding to N2 Ct_LOD_ = 35.0. Equation *P* yields a sum-to-one probability of ‘detected’ or ‘not detected’, wherein the probability was then used as input for a single binomial distribution function to return *N* (e.g., where a high *P* would be likely to return *N* = 1 (i.e., ‘Detected’) and a low *P* would be likely to return *N* = 0 (i.e., ‘Not detected’)) (Figure 1E). Output *N* was then compared with the ground truth (i.e., the clinical test result), which in this case was always ‘detected’, to determine if the simulated result was a true positive or a false negative (Figure 1F). The total number of simulated true positives and false negatives were then used to calculate sensitivity for each epidemiologic week. This was repeated 100 times across all weeks to calculate the average, standard deviation, median, and interquartile range of the simulated sensitivity for each week.

**Figure 1:**
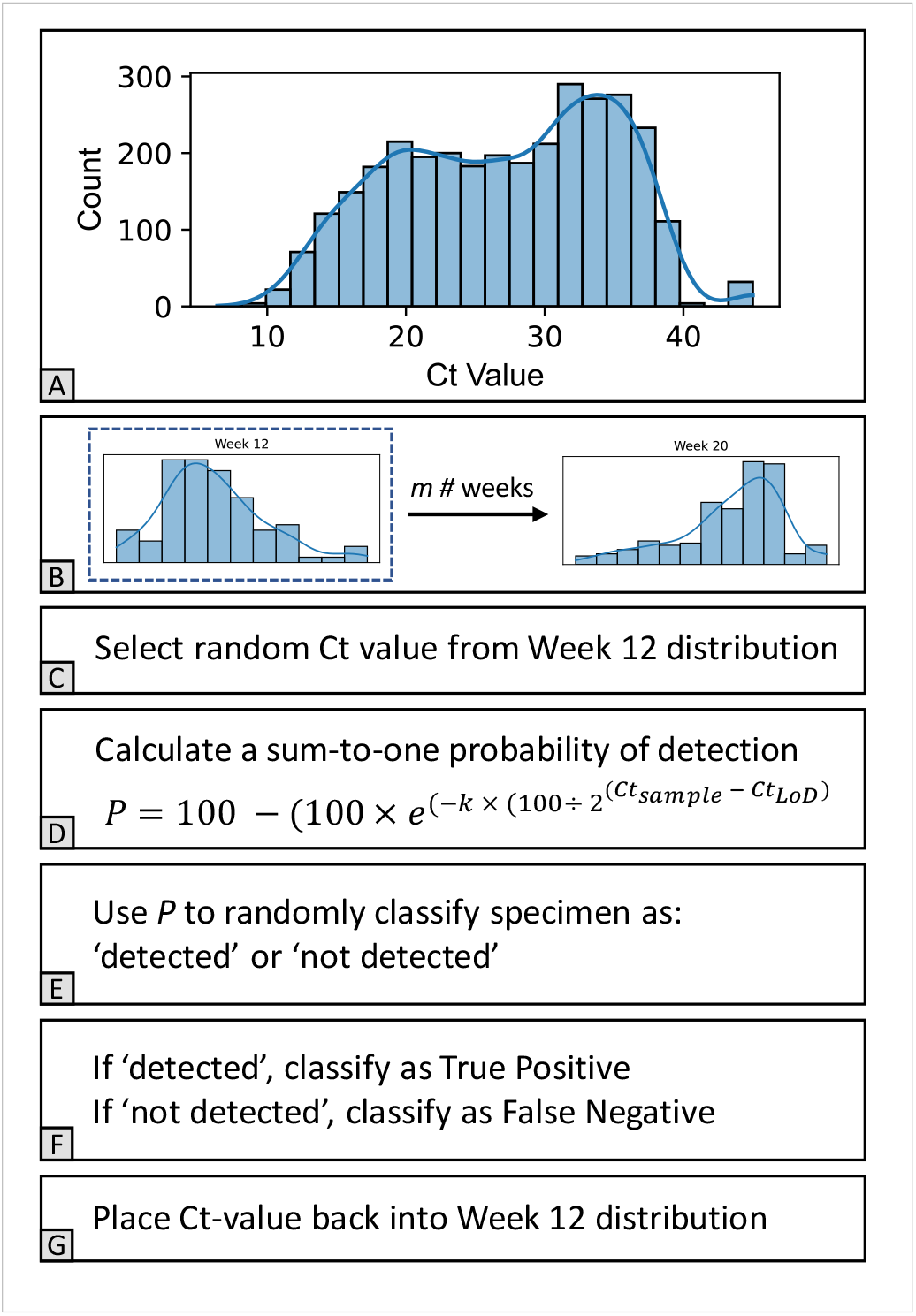
Flow diagram of test performance simulation model. (A) All N2 Ct values belonging to specimens which were clinically reported as ‘detected’ represent the parent distribution. (B) N2 Ct values are separated by epidemiologic week. (C) N2 Ct value is randomly selected from the Week 12 distribution. (D) The selected N2 Ct value is used as input (Ct_sample_) to the probability function to calculate a sum-to-one probability (P). (E) *P* is used to randomly generate a label of ‘detected’ or ‘not detected’. (F) If the randomly generated label is ‘detected’, sample is classified as True Positive. If the randomly generated label is ‘not detected’, sample is classified as False Negative. (G) Sample is placed back into the original epidemiology week distribution. This process repeats 100 times per week across *m* number of epidemiologic weeks for a total of 900 calculations.

A second iteration of simulated sensitivity was carried out using Ct values from the Xpert Xpress SARS-CoV-2 assay, pertaining to the N2 gene target. The LoD for the hypothetical assay for this simulation was defined as the viral concentration corresponding to an N2 Ct value of 40.5, 38.85, and 37.2, which correspond to the LoD of the Xpert assay, and a 0.5- and 1-Log reduction thereof. Ct values were then grouped by epidemiologic weeks which were representative of sample viral burden distributions at our institution during early and late time points during the pandemic. The simulation was repeated 100 times across Ct_LOD_ = 40.5, 38.85, and 37.2, and across selected weeks to calculate the average, standard deviation, median, and interquartile range of the simulated sensitivity for each week.

## RESULTS

### SARS-CoV-2 Testing

The observation period for this project was between March 15, 2020 and July 31, 2020 (corresponding to epidemiologic weeks 12 through 31). The flagship hospital for our HDO is located in New Haven, CT, and we were part of the “first wave” of COVID-19 infections in the USA. We implemented testing locally on March 13, 2020, with the first full week of testing occurring on week 12. COVID-19 positivity rates by PCR testing peaked at week 14 (approx. 30%). This peak has been followed by a long tail of reduced positivity in the setting of dramatically increased outpatient testing. Testing of inpatients and those in the ED has remained stable since early in the pandemic (Figure 2).

**Figure 2:**
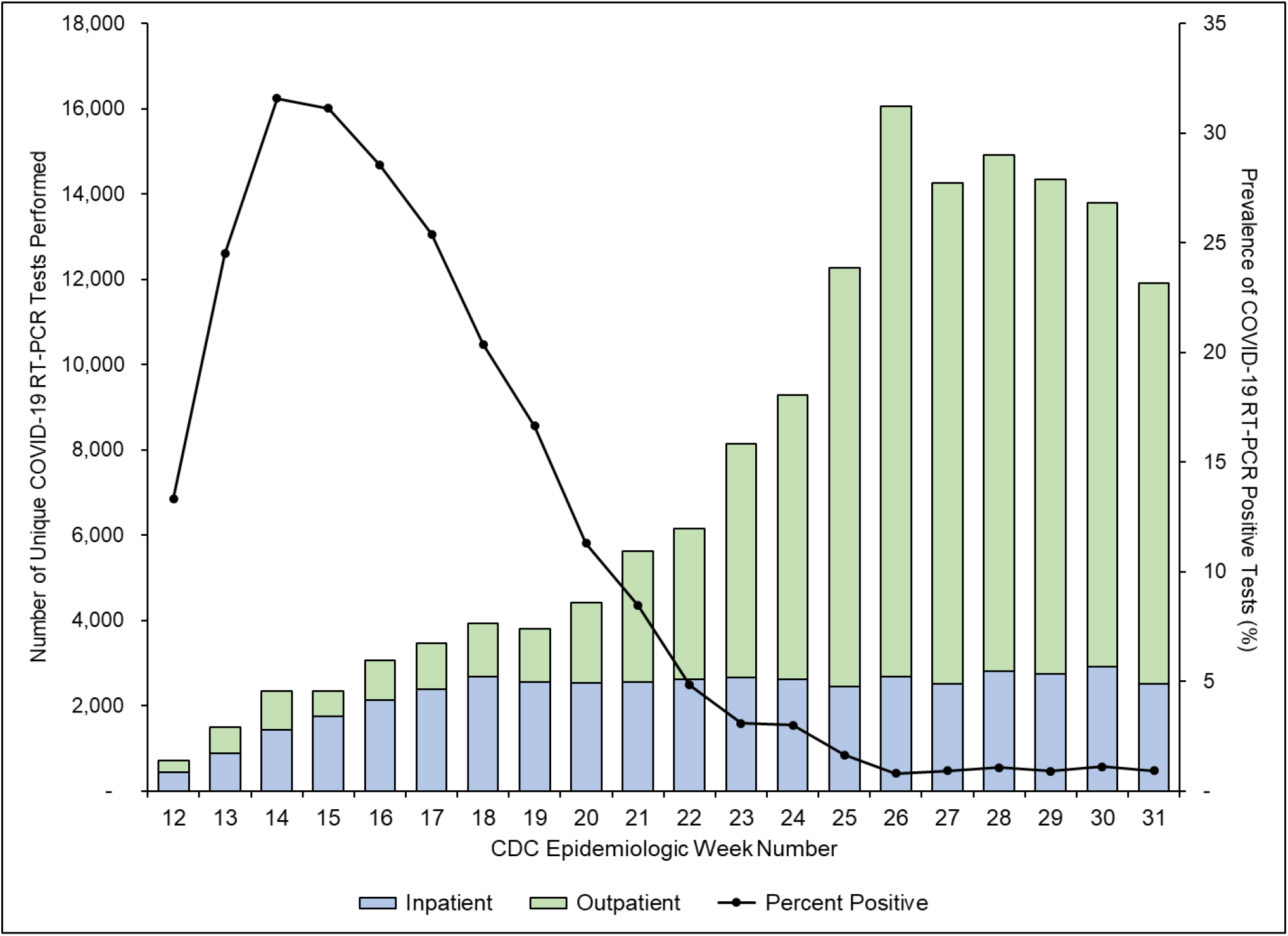
(Left Y-axis) Number of unique SARS-CoV-2 RT-PCR tests grouped by patient location. ‘Inpatient’ represents a summation of samples collected from admitted patients or patients in the emergency department. (Right Y-axis) Number of unique SARS-CoV-2 RT-PCR tests which were ‘positive’. Both data are grouped by CDC epidemiologic week numbers.

### Ct Value Results

While monitoring test performance for quality purposes, we observed that Ct values generated by the available testing platforms, a laboratory developed variation of the CDC assay and the Cepheid GeneXpert Xpress, were increasing with time as the pandemic progressed. We wished to formally characterize this increase by tracking Ct values by week until mid-May 2020 (week 20), a period in which nearly all local testing was performed by one of these two assays. Indeed, Ct values for all gene targets increased with time (Figure 3A – 3D) such that the median Ct values for the N2 gene target increased from 22.7 to 32.8 and 26.0 to 34.8 for the CDC and Xpert assays from week 14 to week 20, respectively. When assessed by linear regression, the slopes for regression lines for all four gene targets were significantly greater than zero. In contrast, Ct values for an influenza A gene target from the Xpert Xpress Influenza A / Influenza B assay observed during the 2019 – 2020 influenza virus season, did not differ as the season progressed, and the slope of the best fit line was not significantly different from zero (Figure 3E)(Supplemental Table 1)(Supplemental Figure 2).

**Figure 3:**
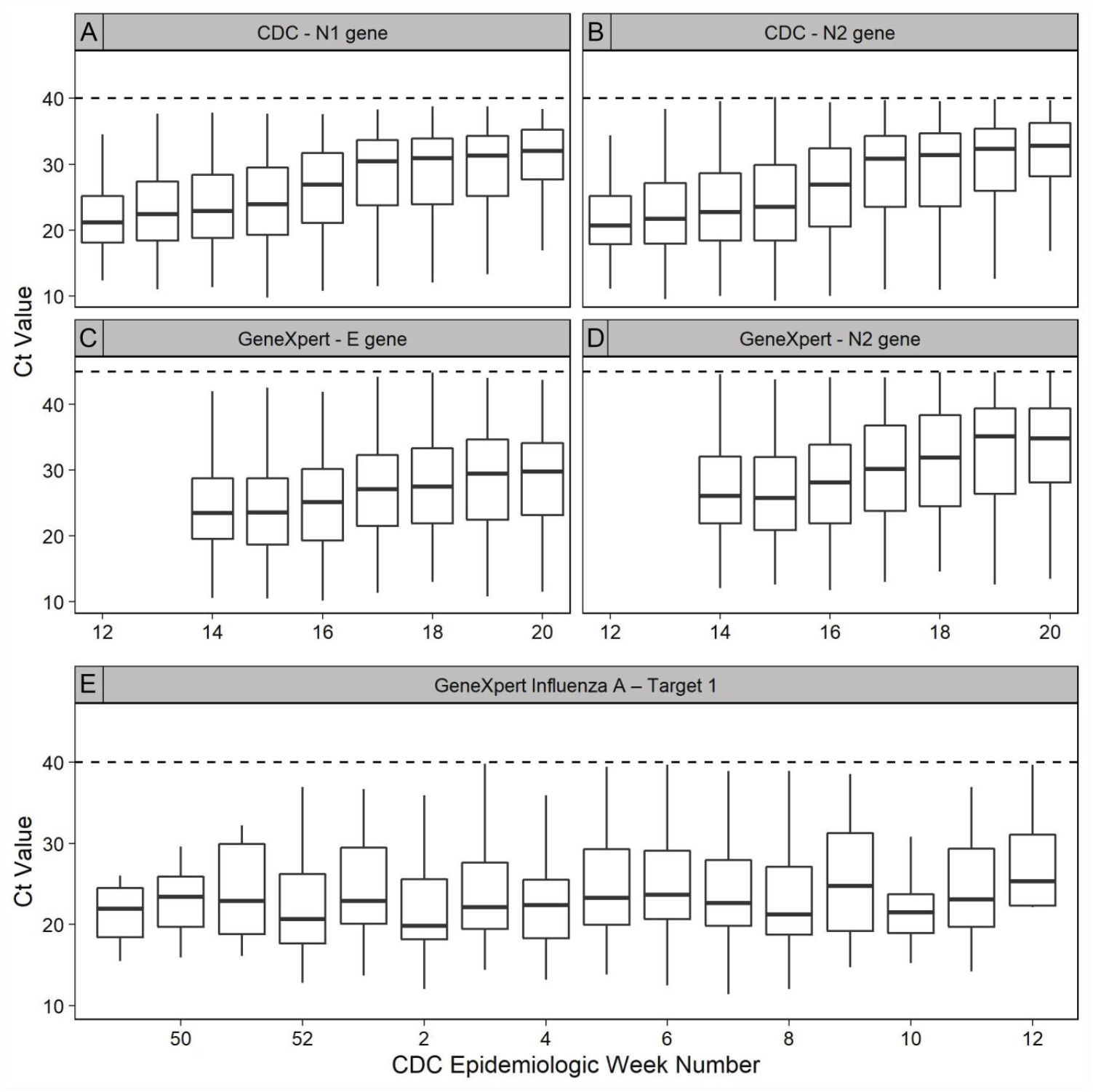
Boxplots representing the Ct value distributions, grouped by epidemiologic week number, test method, and gene target. (A) Modified CDC assay N1 gene target. (B) Modified CDC assay N2 gene target. (C) Cepheid Xpert Xpress SARS-CoV-2 E gene target. (D) Cepheid Xpert Xpress SARS-CoV-2 N2 gene target. (E) Cepheid Xpert Xpress Influenza A gene Target 1. Horizontal dashed lines represent the assay LOD.

### Determination of detection probability as a function of fraction of LoD

We wished to determine the quantitative effect of rising Ct values on diagnostic test performance, but Ct values are not transmutable among assays. Quantitative viral burden measurements would facilitate comparisons among assays, but these are not widely done, and assay limits of detection can be measured in a variety of units that cannot be interconverted. We hypothesized that most extracted RT-PCR assays would have similar behavior at and below the LoD. Additionally, we wished to derive a method wherein the probability of a single sample being reported as positive could be calculated based upon its Ct value compared to the *Ct*_*LoD*_.

We reviewed instructions for use available on the FDA website for commercially available, extracted real-time RT-PCR assays for COVID-19. Data from assays that contained detailed LOD studies, as described in the materials and methods, were extracted, and four assays (Abbott Alinity m, Perkin Elmer, Roche cobas, and Xpert Xpress) met these criteria with individual gene targets from two assays (N and ORF1ab from Perkin Elmer and Target 1 and Target 2 from Roche cobas) being used. Tested concentrations were converted to %-LOD to normalize across testing platforms and units of measure. Plotting %-detected v. %-LoD allowed us to fit an exponential plateau curve (Figure 4) to the data which had an R^2^ = 0.91 and constant (*k*= 0.028737) (Equation A):

**Figure 4:**
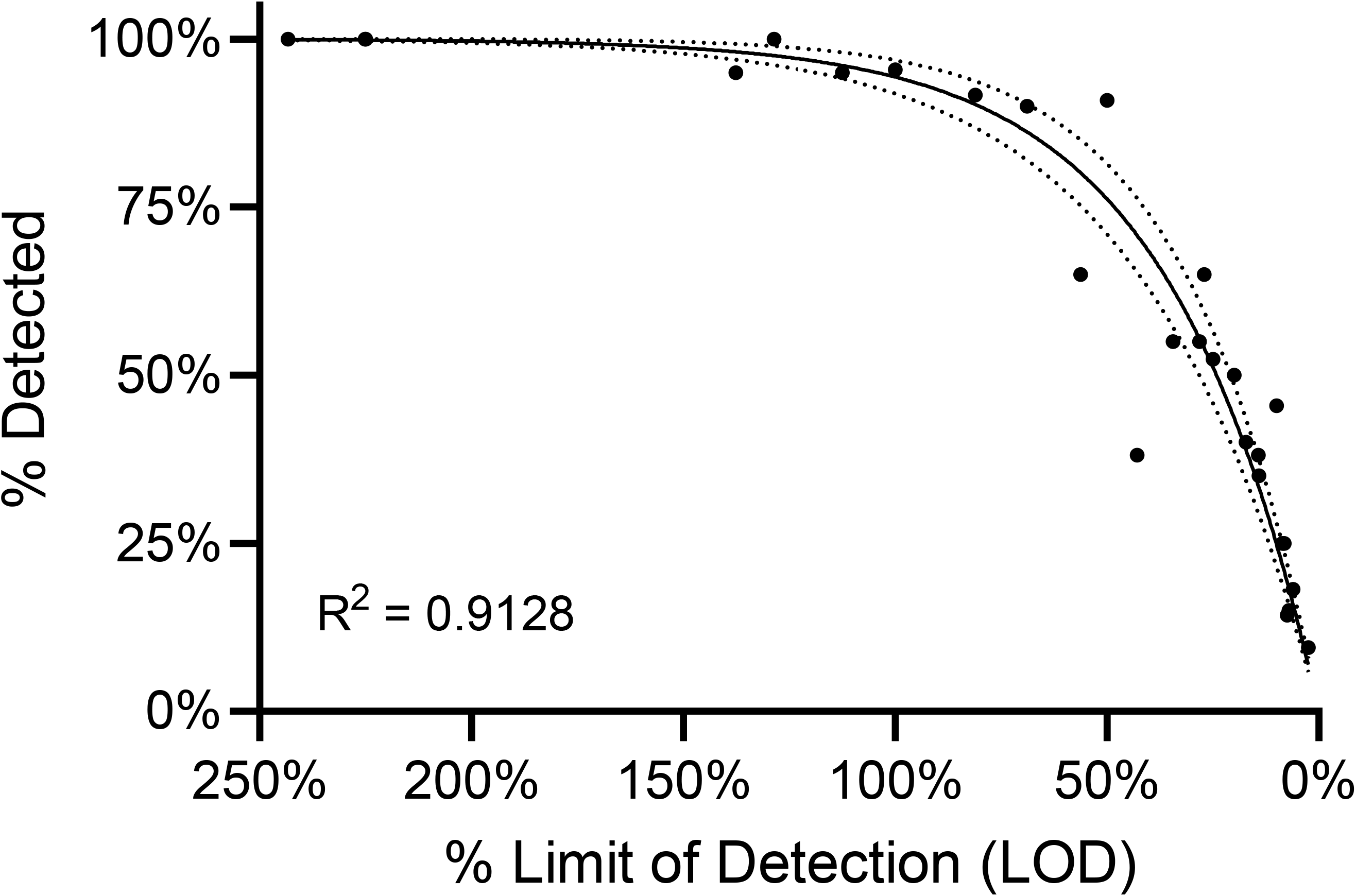
Composite test performance of commercial extracted RT-PCR assays for SARS-CoV-2. Available limit of detection (LOD) data were extracted from publicly available instructions for use (IFU) and normalized to %-LOD as described in methods. The %-Detected at each relative concentration was calculated, plotted, and subjected to non-linear regression with an exponential plateau function with Y0 = 0 and YM = 100%. Best fit line (solid line) and 95% confidence intervals (dashed lines) are shown.

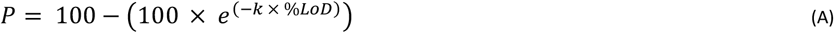

%-LoD can be expressed as relationship between the *Ct*_*sample*_ and *Ct*_*LoD*_ (B):

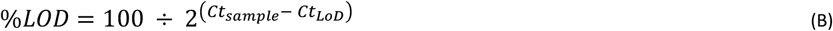

Accordingly, by substituting the %-LoD expression in terms of *Ct*_*LoD*_, equation (C) relates the probability of viral RNA detection by RT-PCR (*P*) as a function of observed Ct value (*Ct*_*sample*_) and *Ct*_*LoD*_, where *k* is a constant.

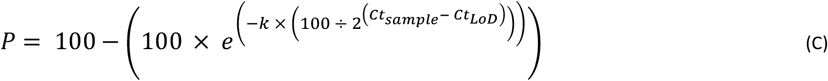

### Test Performance Simulation Results

Using equation (C) which relates the probability (*P*) of detecting a sample based on the relationship of *Ct*_*sample*_ to *Ct*_*LoD*_, we simulated the performance of a single hypothetical assay as observed viral burden among positive samples decreased as the pandemic progressed as summarized in Figure 1. Assuming a relationship between LoD and Ct value is consistent within a given sample, samples with Ct values at the assay LoD should be detected 95% of the time, and samples with Ct values above the *Ct*_*LoD*_ should have a non-zero probability of detection that is inversely related to *Ct*_*sample*_ (i.e., samples with *Ct*_*sample*_ > *Ct*_*LoD*_ may be detected, but less than 95% of the time). Accordingly, for the simulated test performance of a hypothetical assay with a *Ct*_*LoD*_ corresponding to N2 Ct = 35 in the CDC assay, the simulated diagnostic sensitivity decreased from 97.59 (95% CI 97.3 to 97.9) in week 12, to 74.42 (95% CI 73.9 to 75) in week 20. In addition, the coefficient of variation (CV) increased from 1.6% to 3.6% from week 12 to week 20, respectively. At week 16, the simulated diagnostic sensitivity fell below 95% with an average of 94.17 (95% CI 94 to 94.4) (Figure 5).

**Figure 5:**
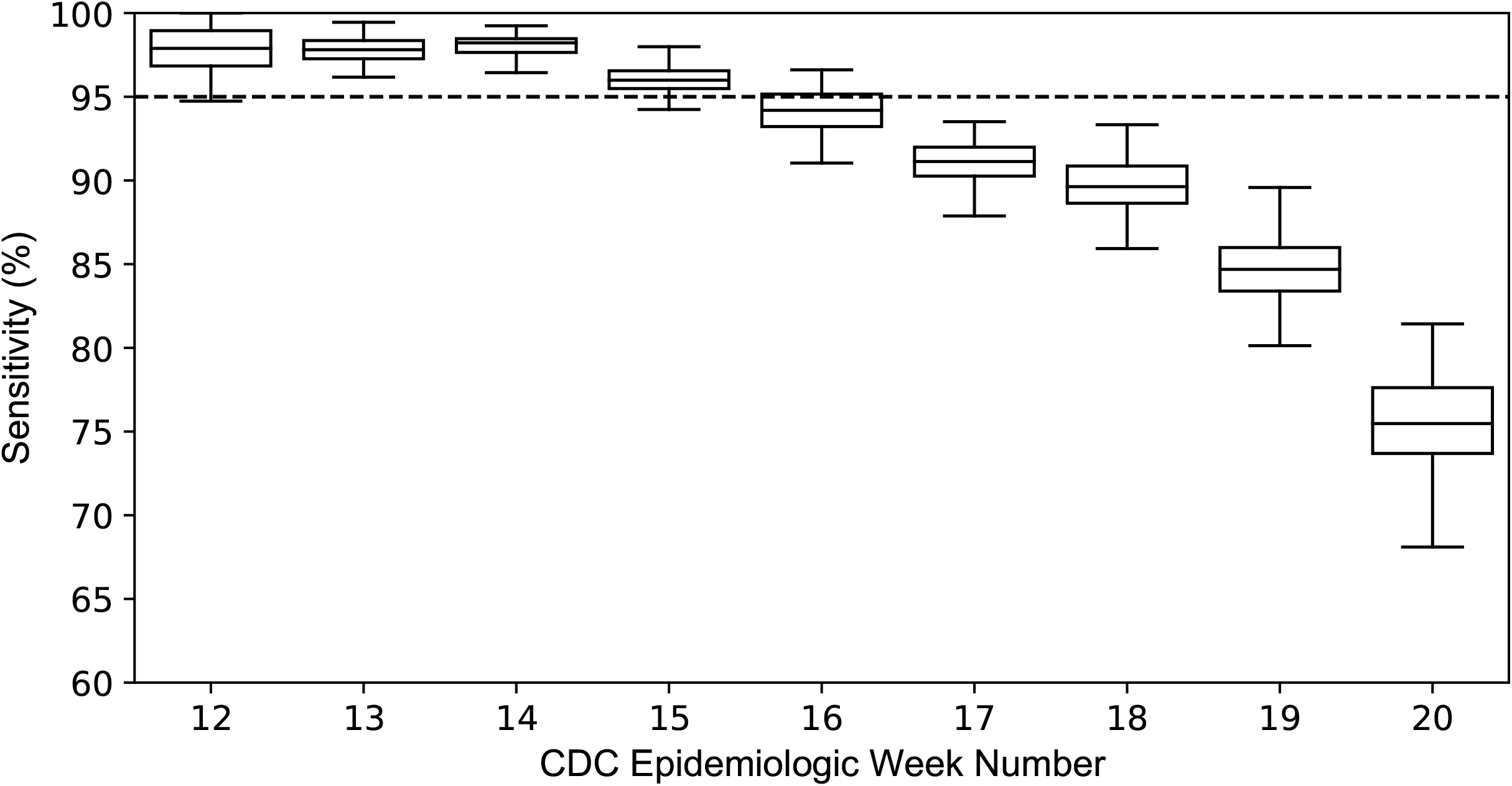
Boxplots representing the distributions of simulated diagnostic sensitivity using Ct values, corresponding to N2 gene target, derived from samples tested by the modified CDC assay. Results are grouped by epidemiologic week number. The horizontal dashed line represents the theoretical LOD probability of detection: 95%.

The second iteration of simulated test performance was carried out using Ct values of the N2 gene target, measured by the Xpert Xpress SARS-CoV-2 assay and then grouped by epidemiologic weeks 14 and 20. The *Ct*_*LoD*_ for the Xpert Xpress assay is available in the IFU, and we were able to simulate the performance of methods with the same relative LoD (*Ct*_*LoD*_ = 40.5) and hypothetical methods with 3-fold and 10-fold reduced LoDs, *Ct*_*LoD*_ = 38.85 and *Ct*_*LoD*_ = 37.2, respectively (Figure 6). The average simulated sensitivity for *Ct*_*LoD*_ = 40.5, 38.85, and 37.20, during epidemiologic week 14 was 98.71 (95% CI 98.5 to 98.9), 97.33 (95% CI 97.1 to 97.5), and 95.79 (95% CI 95.6 to 96), respectively. During week 20, the average sensitivity for *Ct*_*LoD*_ = 40.5, 38.85, and 37.20 was 95.41 (95% CI 95 to 95.8), 85.55 (95% CI 84.9 to 86.2), and 74.38 (95% CI 73.6 to 75.1), respectively (Figure 6).

**Figure 6:**
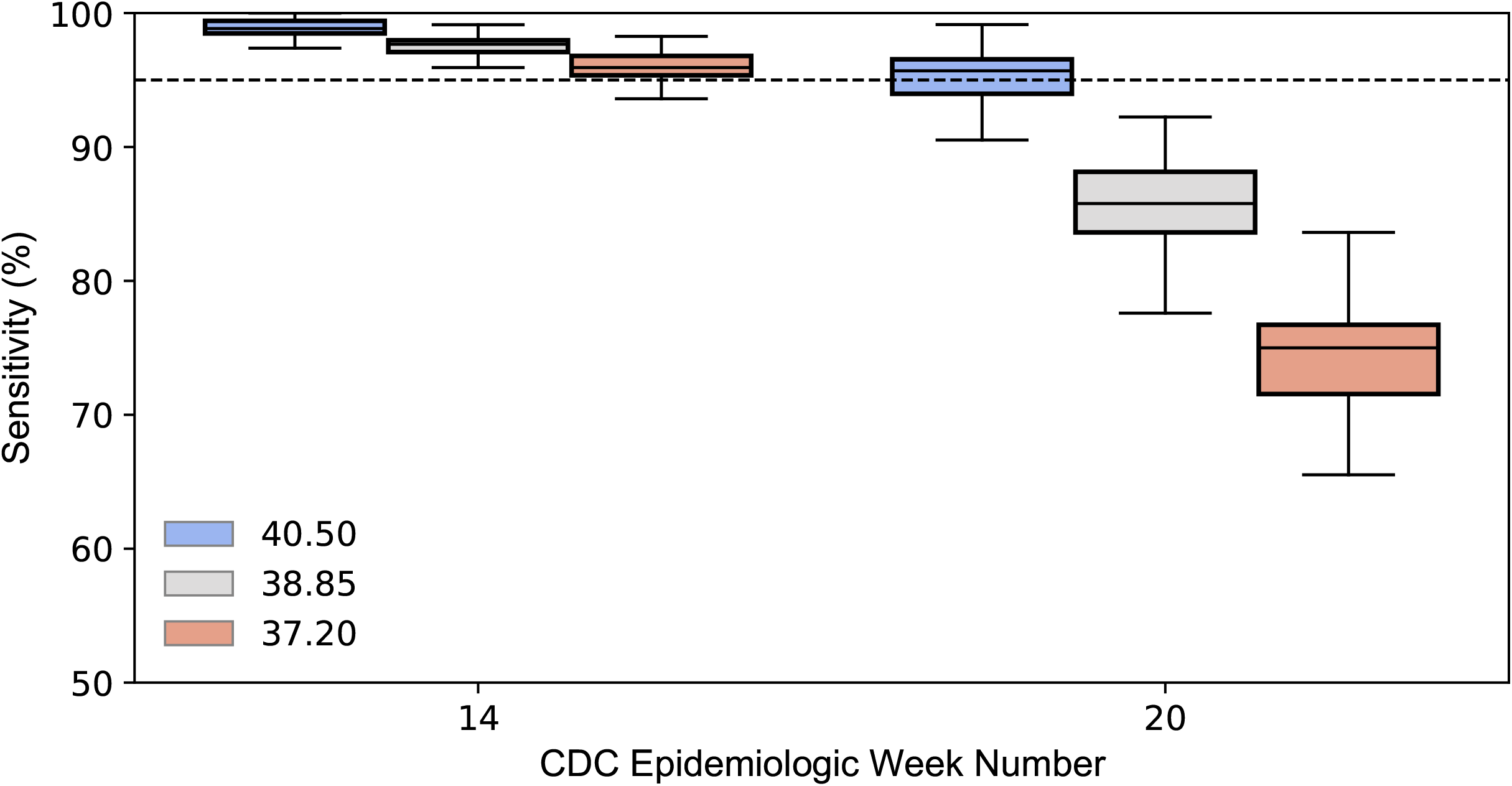
Boxplots representing the distributions of simulated diagnostic sensitivity using Ct values, corresponding to the N2 gene target, derived from samples tested by the Cepheid Xpert Xpress SARS-CoV-2 assay. Results are grouped by epidemiologic week number and *Ct*_*LOD*_ per equation (C). The horizontal dashed line represents the theoretical LOD probability of detection: 95%.

## DISCUSSION

In this report, we describe a novel method for quantitatively describing the effect of variations of sample viral burden on clinical performance characteristics of molecular-based assays. Higher Ct values correlate with lower viral burdens, and lower viral burdens can lead to variable test performance. In most cases where RT-PCR testing is applied for viral diagnostics, viral burden can vary among patients, but we typically operate under the assumption that the relative distribution of viral burdens at any point in time should be relatively constant – i.e., average viral burden shouldn’t vary for patients with influenza in December, January, February or March, all things being equal. It is recognized that different patient populations may have different viral burdens (e.g. adult and pediatric patients for conventional respiratory viruses), but these factors are often controlled or reported in comparative studies of test performance. However, if viral burden varies in a systematic manner that is not controlled for or well-known, test performance characteristics may vary substantially in unexpected ways.

We found that Ct values systematically shifted upward (i.e. toward lower viral burdens) among samples tested within our HDO as the COVID-19 pandemic progressed in Connecticut. Conversely, the distribution of Ct values for influenza testing (Supplemental Table 1 and Supplemental Figure 2), did not change significantly over the course of the 2019 to 2020 flu-season (slope: 0.063 (95% CI −0.02 to 0.23)). We hypothesize that there are two primary reasons for the observed shift in SARS-CoV-2 gene target Ct values: 1) repeat testing of known positive patients later in disease when viral shedding is lower and 2) shifting of testing guidelines to test more patients with less severe symptoms. Repeat testing cannot explain all these changes, as GeneXpert testing at our institution is predominantly performed on newly admitted patients. Indeed, this observation likely warrants further investigation as to the underlying etiology and pathogenesis of SARS-CoV-2. The primary focus of this report, however, was to develop a method to quantitatively describe the effect this observation may have on the diagnostic sensitivity an assay.

The LoD is defined as the concentration at which there is a 95% probability of amplification and detection of target nucleic acid. The second edition of the Clinical and Laboratory Standards Institute (CLSI) guideline EP17-A2 recommends probit analysis, a parametric statistical method for estimating the LoD based on observed detection rates and concomitant nucleic acid concentration data, but probit analysis would not allow us to back calculate discrete probabilities of detection each specimen.(21) Nonetheless, a probit regression was used to model the IFU LoD data which provided a good fit, with an R squared of 0.8767 (Supplemental Figure 1). Instead, we used an exponential method to fit publicly available LoD data to model the probability of detection for Ct values in our dataset. This allowed more flexibility by treating Ct values as continuous input variables for the test performance simulation which could be used to calculate a sum-to-one probability of detection using equation (A). In addition, the non-linear regression resulted in a slightly more optimal R squared value of 0.9128.

We assumed for our model that a 10-fold change in virus concentration would result in a shift of 3.32 cycles across the entire analytical range. While this has been demonstrated for high viral burdens, samples with lower viral burdens near the LoD may not predictably behave in this manner. We also assumed that our baseline tests (e.g. LDT modified CDC assay and Xpert Xpress) were highly sensitive. Our internal comparisons among assays is consistent with this assumption, but more complex assumptions and/or models would be needed to extrapolate the performance of a more sensitive method from the data generated by a less sensitive method. Finally, we did not incorporate test specificity and potential false-positive results into our models. While this could be accomplished through assigning a probability of detection to negative specimens corresponding to a pre-determined false positive rate, it would not have a comparable set of data as that available for test sensitivity.

While our analysis was performed on NP swabs, our modeling data has implications for different viral burdens associated with different specimen types. For example, if viral burdens are 5- to 10-fold lower in alternative specimens compared to NP swabs, then the overall clinical sensitivity of the *same* assay would be expected to vary according to our model. Variations in viral burden with time would compound this change in performance, as well. Indeed, the modeling data presented in this report demonstrate how the rate of detection can vary significantly as the distribution of viral burden changes among samples tested, and this may explain the highly variable performance seen for some sample types such as nasal swabs and saliva.(17, 23)

In order to interpret the effects of samples with lower viral burdens on test performance characteristics in publications, whether in comparisons between methods or sample types, rigorous reporting of study parameters as detailed in the STARD 2015 guidelines is necessary.(24) Moreover, compared to other studies of viral diagnostics, it is even more important for COVID studies to detail the provenance of the specimens used including dates of sample collection, explicit case-finding criteria, and local positivity rates. Currently, the literature predominantly reports on samples collected in late March to early May, when testing of asymptomatic patients was not as prevalent.(2, 3, 25) As the publication cycle progresses and studies from later months emerge, it will be important to observe for greater variation in performance characteristics as predicted by and demonstrated in this report. The Ct values of patient specimens are important data to contextualize study findings. However, Ct ranges alone are not sufficient due to the stochastic nature of detection near the LoD, and the proportion of specimens with a low viral burden as evidenced by higher Ct values is an important metric rather than simply the maximum observed Ct.(7) Subsequently, concordance and/or percent-positive-agreement analysis should be tied to ranges of Ct values from well-studied assays in order to properly assess assay performance on different patient populations. Broadly, these parameters will help guide the interpretation and applicability of study results for different institutions and potentially different clinical use case scenarios.

More fundamentally, variation in assay performance over time may be partly attributable to changes in case definition. Whereas early in the pandemic case definitions included clinical criteria when testing was more restricted, widespread testing of asymptomatic patients has led to laboratory test-based case definitions. In lieu of a definitive diagnostic gold standard, high sensitivity methods have been used either individually or with other high sensitivity methods as part of a composite reference standard. Importantly, a single result arising from the use of only one high-sensitivity method as a reference standard may be limited by the stochastic nature of test performance near the LoD. Multi-target assays are likely to be less susceptible to stochastic effects than single-target assays, but also introduce the need for studies to precisely define how each target result is translated into an overall positive or negative specimen result. Though marked discordance between high sensitivity methods at high Ct values has not been observed in several studies, discordance may emerge in data from later months in the pandemic.(2, 3, 7) These considerations again highlight the need to evaluate high sensitivity methods over time, individually and together, in order to capture variations in diagnostic sensitivity.

Laboratory testing, particularly molecular-based assays, are often thought of being deterministic in nature when generating a result – i.e., if the RNA is present, it will be detected. Accordingly, if the test methodology is deterministic and stationary, we often think of test performance as stationary. However, as viral burden in the sample approaches the LoD, it is known that these tests exhibit more stochastic characteristics. At low concentrations of pathogen nucleic acid, chance can dictate whether target nucleic acid is captured in a pipet, bound during the extraction phase, efficiently eluted, or pipetted into the final reaction mixture. All of this complexity is combined to determine the probability of detection at and below the assay LoD. These data demonstrate how variations in characteristics of the underlying test population should be considered when considering the static nature or dynamic nature of test performance.

## Data Availability

Not available.

## Abbreviations

COVID-19: Coronavirus disease of 2019
Ct value: Cycle-threshold value
EHR: Electronic health record
EUA: Emergency use authorization
HDO: Healthcare delivery organization
IFU: Instructions for use
IVD: In-vitro diagnostics
KS_d_: Kolmogorov–Smirnov distance
LIS: Laboratory information system
LOD: Limit of detection
NAAT: nucleic acid amplification tests
NPS: Nasopharyngeal swab
RT-PCR: Real-time reverse transcription-PCR
SARS-CoV-2: Severe acute respiratory syndrome coronavirus 2
UTM: universal transport medium
VTM: viral transport medium
WHVA: West Haven, Veteran’s Administration Hospital

## FIGURE LEGENDS

**Supplemental Table 1:**
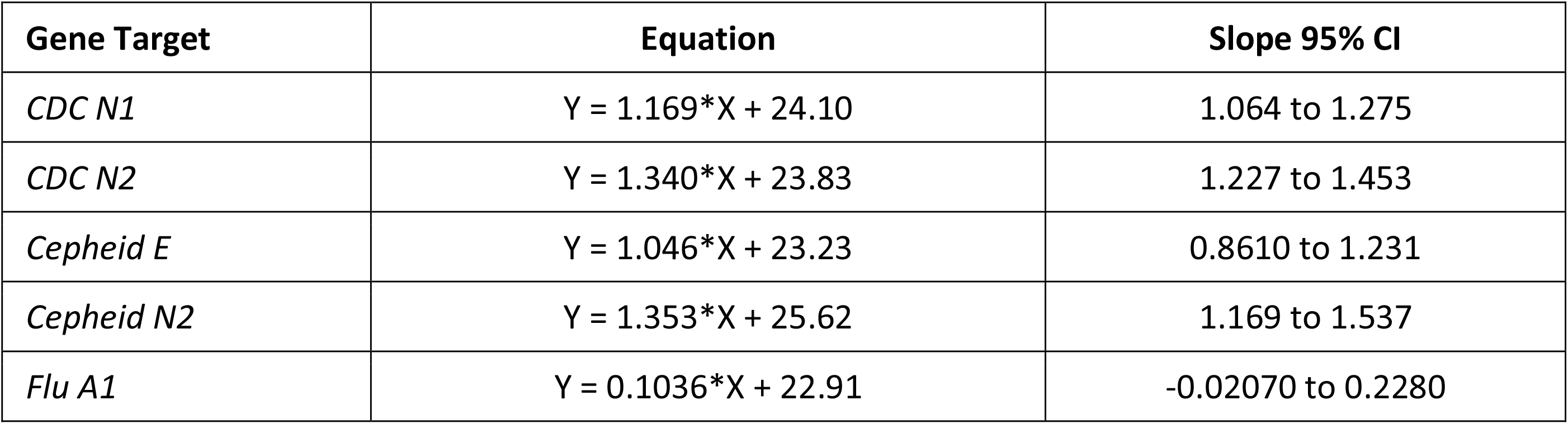
Line of best fit equations and the associated 95% confidence intervals for the slope. Derived from a simple linear regression performed on the Ct values measured from ‘positive’ samples and the associated gene targets.

| Gene Target | Equation | Slope 95% CI |
| --- | --- | --- |
| *CDC N1* | $Y = 1.169 * X + 24.10$ | 1.064 to 1.275 |
| *CDC N2* | $Y = 1.340 * X + 23.83$ | 1.227 to 1.453 |
| *Cepheid E* | $Y = 1.046 * X + 23.23$ | 0.8610 to 1.231 |
| *Cepheid N2* | $Y = 1.353 * X + 25.62$ | 1.169 to 1.537 |
| *Flu A1* | $Y = 0.1036 * X + 22.91$ | -0.02070 to 0.2280 |

**Supplemental Table 2:** Data extracted from commercial assay instructions for use. Roche cobas – Target 2 concentration 0.003 TCID50/mL of 0.003 TCID50/mL was excluded based on outlier analysis.

| Assay | LOD | LOD Measure | Conc Tested | %LOD | Detected | Tested | % Pos |
| --- | --- | --- | --- | --- | --- | --- | --- |
| Alinity M | 0.0037 | TCID50/mL | 0.009 | 243.2% | 24 | 24 | 100.0% |
|  |  |  | 0.003 | 81.1% | 22 | 24 | 91.7% |
|  |  |  | 0.001 | 27.0% | 13 | 20 | 65.0% |
|  |  |  | 0.0003 | 8.1% | 6 | 24 | 25.0% |
| Perkin Elmer (N Gene) | 24.9 | cp/mL | 34.25 | 137.6% | 19 | 20 | 95.0% |
|  |  |  | 17.13 | 68.8% | 18 | 20 | 90.0% |
|  |  |  | 8.56 | 34.4% | 11 | 20 | 55.0% |
|  |  |  | 4.28 | 17.2% | 8 | 20 | 40.0% |
|  |  |  | 2.14 | 8.6% | 5 | 20 | 25.0% |
| Perkin Elmer (ORF1ab) | 9.3 | cp/mL | 20.93 | 225.1% | 20 | 20 | 100.0% |
|  |  |  | 10.46 | 112.5% | 19 | 20 | 95.0% |
|  |  |  | 5.23 | 56.2% | 13 | 20 | 65.0% |
|  |  |  | 2.62 | 28.2% | 11 | 20 | 55.0% |
|  |  |  | 1.31 | 14.1% | 7 | 20 | 35.0% |
|  |  |  | 0.65 | 7.0% | 3 | 20 | 15.0% |
| Roche Cobas (Target 1) | 0.007 | TCID50/mL | 0.009 | 128.6% | 21 | 21 | 100.0% |
|  |  |  | 0.003 | 42.9% | 8 | 21 | 38.1% |
|  |  |  | 0.001 | 14.3% | 8 | 21 | 38.1% |
| Roche Cobas (Target 2) | 0.004 | TCID50/mL | 0.009 | 225.0% | 21 | 21 | 100.0% |
|  |  |  | 0.001 | 25.0% | 11 | 21 | 52.4% |
|  |  |  | 0.0003 | 7.5% | 3 | 21 | 14.3% |
|  |  |  | 0.0001 | 2.5% | 2 | 21 | 9.5% |
| Xpert Xpress | 0.005 | PFU/mL | 0.005 | 100.0% | 21 | 22 | 95.5% |
|  |  |  | 0.0025 | 50.0% | 20 | 22 | 90.9% |
|  |  |  | 0.001 | 20.0% | 11 | 22 | 50.0% |
|  |  |  | 0.0005 | 10.0% | 10 | 22 | 45.5% |
|  |  |  | 0.0003 | 6.0% | 4 | 22 | 18.2% |

Abbreviations: cp = Copies; LOD = Limit of detection; mL = milliliter; PFU = Plaque forming units; Pos = Positive; TCID50 = Median Tissue Culture Infectious Dose;

**Supplemental Figure 1:**
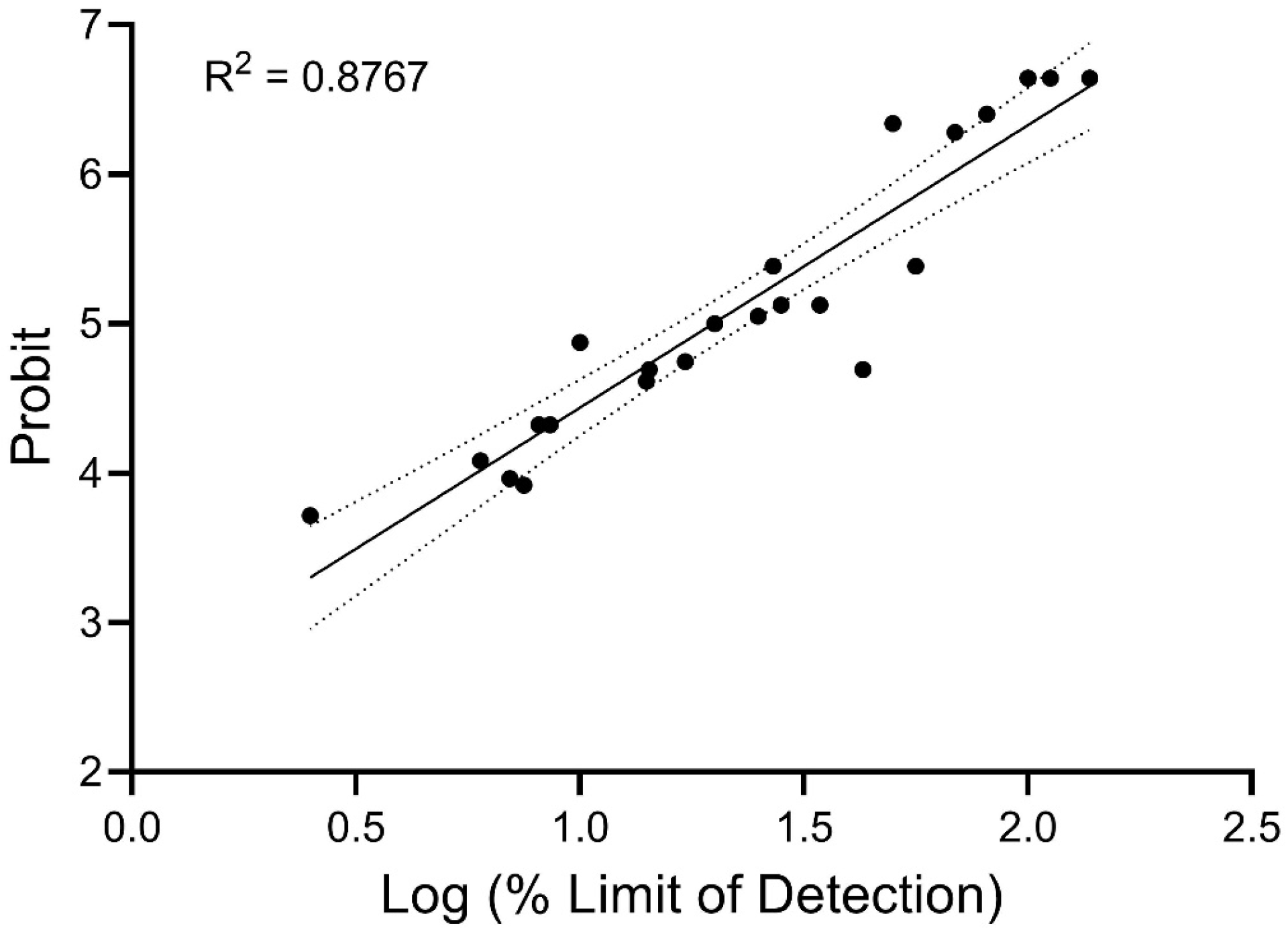
Composite test performance of commercial extracted RT-PCR assays for SARS-CoV-2. Available limit of detection (LOD) data were extracted from publicly available instructions for use (IFU) and normalized to Log %-LOD as described in methods and plotted against probit units determined using GraphPad Prism v8. Best fit line (solid line) and 95% confidence intervals (dashed lines) are shown.

**Supplemental Figure 2:**
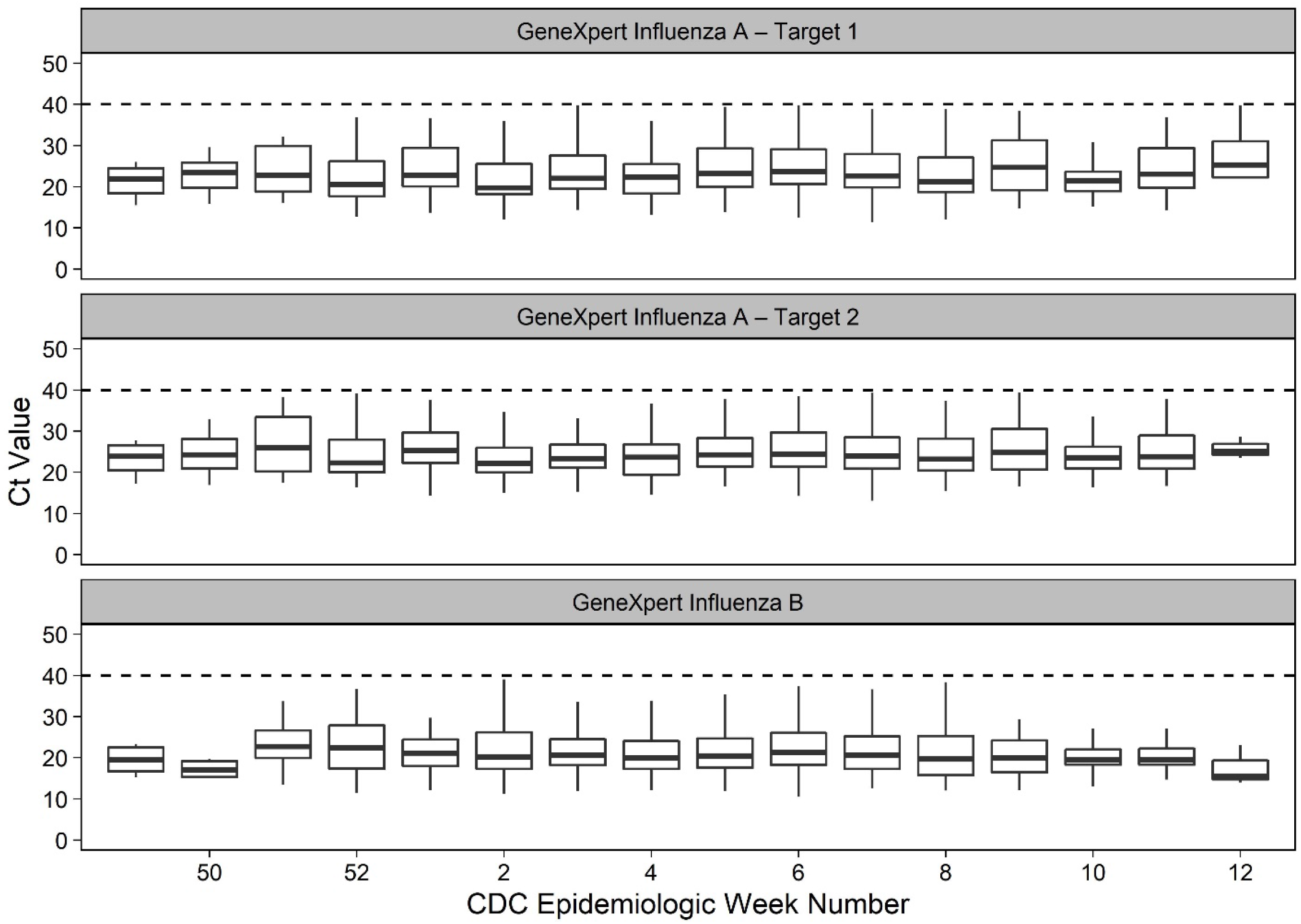
Boxplots representing the Ct value distributions for Ct values derived from the Cepheid Xpert Xpress Influenza A / Influenza B assay during flu-season 2019 to 2020. Results are grouped by epidemiologic week number and gene target. Horizontal dashed lines represent the assay LOD.

## BIBLIOGRAPHY

1. Fung B, Gopez A, Servellita V, Arevalo S, Ho C, Deucher A, Thornborrow E, Chiu C, Miller S. 2020. Direct Comparison of SARS-CoV-2 Analytical Limits of Detection across Seven Molecular Assays. J Clin Microbiol 58.

2. Moran A, Beavis KG, Matushek SM, Ciaglia C, Francois N, Tesic V, Love N. 2020. Detection of SARS-CoV-2 by Use of the Cepheid Xpert Xpress SARS-CoV-2 and Roche cobas SARS-CoV-2 Assays. J Clin Microbiol 58.

3. Broder K, Babiker A, Myers C, White T, Jones H, Cardella J, Burd EM, Hill CE, Kraft CS. 2020. Test Agreement between Roche Cobas 6800 and Cepheid GeneXpert Xpress SARS-CoV-2 Assays at High Cycle Threshold Ranges. J Clin Microbiol 58.

4. Liotti FM, Menchinelli G, Marchetti S, Morandotti GA, Sanguinetti M, Posteraro B, Cattani P. 2020. Evaluation of three commercial assays for SARS-CoV-2 molecular detection in upper respiratory tract samples. Eur J Clin Microbiol Infect Dis.

5. Rhoads DD, Cherian SS, Roman K, Stempak LM, Schmotzer CL, Sadri N. 2020. Comparison of Abbott ID Now, DiaSorin Simplexa, and CDC FDA Emergency Use Authorization Methods for the Detection of SARS-CoV-2 from Nasopharyngeal and Nasal Swabs from Individuals Diagnosed with COVID-19. J Clin Microbiol 58.

6. Craney AR, Velu PD, Satlin MJ, Fauntleroy KA, Callan K, Robertson A, La Spina M, Lei B, Chen A, Alston T, Rozman A, Loda M, Rennert H, Cushing M, Westblade LF. 2020. Comparison of Two High-Throughput Reverse Transcription-PCR Systems for the Detection of Severe Acute Respiratory Syndrome Coronavirus 2. J Clin Microbiol 58.

7. Smithgall MC, Scherberkova I, Whittier S, Green DA. 2020. Comparison of Cepheid Xpert Xpress and Abbott ID Now to Roche cobas for the Rapid Detection of SARS-CoV-2. J Clin Virol 128:104428.

8. Zhen W, Manji R, Smith E, Berry GJ. 2020. Comparison of Four Molecular In Vitro Diagnostic Assays for the Detection of SARS-CoV-2 in Nasopharyngeal Specimens. J Clin Microbiol 58.

9. Tanida K, Koste L, Koenig C, Wenzel W, Fritsch A, Frickmann H. 2020. Evaluation of the automated cartridge-based ARIES SARS-CoV-2 Assay (RUO) against automated Cepheid Xpert Xpress SARS-CoV-2 PCR as gold standard. Eur J Microbiol Immunol (Bp).

10. Green DA, Zucker J, Westblade LF, Whittier S, Rennert H, Velu P, Craney A, Cushing M, Liu D, Sobieszczyk ME, Boehme AK, Sepulveda JL. 2020. Clinical Performance of SARS-CoV-2 Molecular Tests. J Clin Microbiol 58.

11. Moore NM, Li H, Schejbal D, Lindsley J, Hayden MK. 2020. Comparison of Two Commercial Molecular Tests and a Laboratory-Developed Modification of the CDC 2019-nCoV Reverse Transcriptase PCR Assay for the Detection of SARS-CoV-2. J Clin Microbiol 58.

12. Dust K, Hedley A, Nichol K, Stein D, Adam H, Karlowsky JA, Bullard J, Van Caeseele P, Alexander DC. 2020. Comparison of Commercial Assays and Laboratory Developed Tests for Detection of SARS-CoV-2. J Virol Methods 113970.

13. Lieberman JA, Pepper G, Naccache SN, Huang M-L, Jerome KR, Greninger AL. 2020. Comparison of Commercially Available and Laboratory-Developed Assays for In Vitro Detection of SARS-CoV-2 in Clinical Laboratories. J Clin Microbiol 58.

14. Jin R, Pettengill MA, Hartnett NL, Auerbach HE, Peiper SC, Wang Z. 2020. Commercial SARS-CoV-2 Molecular Assays: Superior Analytical Sensitivity of cobas SARS-CoV-2 Relative to NxTAG Cov Extended Panel and ID NOW COVID-19 Test. Arch Pathol Lab Med.

15. Hanson KE, Caliendo AM, Arias CA, Englund JA, Lee MJ, Loeb M, Patel R, El Alayli A, Kalot MA, Falck-Ytter Y, Lavergne V, Morgan RL, Murad MH, Sultan S, Bhimraj A, Mustafa RA. 2020. Infectious Diseases Society of America Guidelines on the Diagnosis of COVID-19. Clin Infect Dis.

16. McCormick-Baw C, Morgan K, Gaffney D, Cazares Y, Jaworski K, Byrd A, Molberg K, Cavuoti D. 2020. Saliva as an Alternate Specimen Source for Detection of SARS-CoV-2 in Symptomatic Patients Using Cepheid Xpert Xpress SARS-CoV-2. J Clin Microbiol 58.

17. Wyllie AL, Fournier J, Casanovas-Massana A, Campbell M, Tokuyama M, Vijayakumar P, Warren JL, Geng B, Muenker MC, Moore AJ, Vogels CBF, Petrone ME, Ott IM, Lu P, Venkataraman A, Lu-Culligan A, Klein J, Earnest R, Simonov M, Datta R, Handoko R, Naushad N, Sewanan LR, Valdez J, White EB, Lapidus S, Kalinich CC, Jiang X, Kim DJ, Kudo E, Linehan M, Mao T, Moriyama M, Oh JE, Park A, Silva J, Song E, Takahashi T, Taura M, Weizman O-E, Wong P, Yang Y, Bermejo S, Odio CD, Omer SB, Dela Cruz CS, Farhadian S, Martinello RA, Iwasaki A, Grubaugh ND, Ko AI. 2020. Saliva or Nasopharyngeal Swab Specimens for Detection of SARS-CoV-2. N Engl J Med 383:1283–1286.

18. Hou H, Chen J, Wang Y, Lu Y, Zhu Y, Zhang B, Wang F, Mao L, Tang Y-W, Hu B, Ren Y, Sun Z. 2020. Multicenter Evaluation of the Cepheid Xpert Xpress SARS-CoV-2 Assay for the Detection of SARS-CoV-2 in Oropharyngeal Swab Specimens. J Clin Microbiol 58.

19. Bland JM, Altman DG. 2015. Statistics Notes: Bootstrap resampling methods. BMJ 350:h2622.

20. Massey FJ. 1951. The Kolmogorov-Smirnov Test for Goodness of Fit. J Am Stat Assoc 46:68–78.

21. Clsi. 2012. Evaluation of detection capability for clinical laboratory measurement procedures; approved guideline—second edition. CLSI document EP17-A2.

22. Cradic K, Lockhart M, Ozbolt P, Fatica L, Landon L, Lieber M, Yang D, Swickard J, Wongchaowart N, Fuhrman S, Antonara S. 2020. Clinical Evaluation and Utilization of Multiple Molecular In Vitro Diagnostic Assays for the Detection of SARS-CoV-2. Am J Clin Pathol 154:201–207.

23. Callahan C, Lee R, Lee G, Zulauf KE, Kirby JE, Arnaout R. 2020. Nasal-Swab Testing Misses Patients with Low SARS-CoV-2 Viral Loads. medRxiv.

24. Bossuyt PM, Reitsma JB, Bruns DE, Gatsonis CA, Glasziou PP, Irwig L, Lijmer JG, Moher D, Rennie D, de Vet HCW, Kressel HY, Rifai N, Golub RM, Altman DG, Hooft L, Korevaar DA, Cohen JF, STARD Group. 2015. STARD 2015: an updated list of essential items for reporting diagnostic accuracy studies. Clin Chem 61:1446–1452.

25. Tu Y-P, Jennings R, Hart B, Cangelosi GA, Wood RC, Wehber K, Verma P, Vojta D, Berke EM. 2020. Swabs Collected by Patients or Health Care Workers for SARS-CoV-2 Testing. N Engl J Med 383:494–496.

